# Infectivity and persistence of influenza viruses in raw milk

**DOI:** 10.1101/2024.10.10.24315269

**Authors:** Alessandro Zulli, Mengyang Zhang, Sehee Jong, Catherine Blish, Alexandria B. Boehm

**Affiliations:** Department of Civil and Environmental Engineering, Stanford University, 473 Via Ortega, Stanford, California, 94305; Department of Medicine, Stanford University School of Medicine, Stanford, CA 94305, United States

**Keywords:** influenza A, milk, persistence, infectivity, avian influenza

## Abstract

Influenza A viruses present a significant public health risk, with recent outbreaks of highly pathogenic avian influenza (HPAI) H5N1 in dairy cattle raising concerns about potential transmission through raw milk consumption. This study investigated the persistence of influenza A virus PR8 (IAV PR8) in raw cow milk at 4 °C. We found that IAV PR8 remained infectious in raw milk for up to 5 days, with a decay rate constant of −2.05 day^−1^. In contrast, viral RNA remained detectable and stable for at least 57 days, with no significant degradation. Pasteurization (63°C for 30 minutes) significantly reduced detectable viral RNA concentrations, but reduction was less than 1 log. These findings highlight the potential risk of zoonotic virus transmission through raw milk consumption and underscore the importance of milk pasteurization. The prolonged persistence of viral RNA in both raw and pasteurized milk has implications for food safety assessments and environmental monitoring, particularly in the context of environmental surveillance for influenza viruses.

**Synopsis:** Influenza A RNA is persistent in milk, even after pasteurization, and it remains infectious for 5 days in refrigerated milk.

## Introduction

Influenza A remains one of the leading causes of mortality and morbidity worldwide, with the World Health Organization estimating that it causes up to 650,000 deaths and 1 billion cases each year.^1^ In the United States, it is responsible for an estimated annual 41 million infections, and 51,000 deaths, infecting approximately 8% of the US population.^2,3^ In addition, it is estimated to be responsible for $11.2 billion dollars in economic losses each year within the United States.^4,5^

Influenza A has demonstrated high rates of mutation and zoonotic potential, making it difficult to take appropriate preventative measures.^6,7^ The zoonotic potential of influenza A viruses is best demonstrated by the first influenza pandemic of the 21st century, the 2009 H1N1 virus, commonly known as “swine flu”.^8^ This outbreak is estimated to have been responsible for 700 million to 1.4 billion infections in the 2009-2010 season.^9^ As its name implies, genetic analysis of the 2009 H1N1 virus revealed its origin to be reassortment and subsequent zoonotic transmission from swine to humans.^8,10,11^ Highly pathogenic avian influenza H5N1 (HPAI), has demonstrated similar reassortment and zoonotic transmission to the 2009 H1N1 virus, and has recently been cause for significant concern in the United States and worldwide.^7,12^ Specifically, H5N1 clade 2.3.4.4 has affected over 100 million poultry since 2022, and, more recently, 194 herds of cattle, having made the interspecies jump.^13–17^ Prior to 2024, H5N1 infection in cows had not been documented.^15^

Interestingly, infectious H5N1 is shed into milk of infected, lactating cows.^17^ Several studies have shown pasteurization is effective in inactivating influenza virus in milk, however, consumption of raw (i.e., unpasteurized) milk is common.^18^ The latest United States Department of Agriculture survey from 2019 reported 4.4% of Americans consume raw milk at least once a year, and 1.6% (approximately 5 million people) report consuming raw milk once a month. We reviewed the literature on October 7, 2024 using keywords “influenza raw milk”, “influenza pasteurization” and “infectivity influenza raw milk”. We identified no studies documented persistence of infective viruses in raw milk, and limited studies documenting the effect of pasteurization of influenza A infectivity in raw milk, and two of these have not been peer-reviewed (pre-prints).^18–20^ Given the high consumption of raw milk in the United States, and increasing trends in outbreaks associated with raw milk consumption, understanding the persistence of infectious influenza A virus in milk could be particularly useful to better understanding infection risks associated with this environmental exposure route.^21–23^

Genetic material of H5N1 has been detected in pasteurized milk products across the United States.^19,20,24^ The presence of H5N1 RNA in milk products has been linked to its appearance in wastewater in a number of states, and served to alert public health officials to the potential of nearby infected cattle herds.^14,25^ Our literature review identified no papers examining the effect of pasteurization on influenza A viral RNA in milk; such information will be useful in advancing the use of wastewater for tracking the spread of H5N1.^26,27^

In this study, we used influenza A H1N1, to assess the persistence of influenza A virus in raw milk. Our results demonstrate that IAV PR8 remains infectious in raw milk for up to 5 days at 4 °C, while their genetic material remains detectable in raw milk for 8 weeks, showing little to no degradation at 4 °C. Lastly, the RNA of H1N1 showed significant degradation after heating at 63 °C for 30 min, which represents the standard process used for milk pasteurization.

## Methods

### Virus propagation and purification

IAV PR8 was propagated in Madin Darby canine kidney (MDCK) cells.^28^ MDCK cells (CCL-34, ATCC) were purchased from ATCC and IAV PR8 was kindly provided by Dr. Jeffrey Glenn. Details of virus propagation are provided in the supporting information (SI).

IAV PR8 virus stock was purified and concentrated using ultracentrifugation with 30% sucrose cushion.^29^ Briefly, the crude virus suspension collected from the infected MDCK cells was centrifuged at 4,000 g for 15 minutes to remove cell debris before ultracentrifugation. Then 180 mL of the supernatant was processed by ultracentrifugation with a 30% of sucrose cushion at 112,400 g for 90 minutes at 4 °C. The IAV PR8 virus pellets were resuspended in 1 mL of PBS buffer and stored at −80 °C until use.

### Influenza A virus persistence in raw milk

Raw milk was collected from a local dairy farm in California and stored at 4 °C until persistence experiments (used within 24 hours). Purified IAV PR8 was spiked into the raw milk at the ratio of 1:10 to achieve final IAV PR8 concentrations of around 10^5^-10^6^ TCID_50_/mL. The mixture of IAV PR8 and raw milk was incubated at 4 °C for 57 days. The experiment was performed in triplicate. During the incubation period, at day 0, 1, 3 and 5, 60 µL of the sample was withdrawn from each experiment for 50% tissue culture infectious dose (TCID_50_) assay to measure the concentration of viable IAV PR8; at day 0, 1, 3, 5, 7, 21, 28, and 35, 60 µL of the sample was withdrawn from each tube for RNA extraction to measure the concentration of IAV PR8 RNA. TCID_50_ assay and RNA extraction was performed immediately after the samples were withdrawn.

The decay rate constant of viable IAV PR8 (*k*_*infectivity*_)was estimated by fitting a first-order decay model to the corresponding virus infectivity data following equation (1):

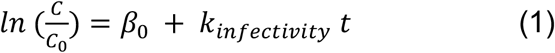

where *C* is the average concentration of viable IAV PR8 measured at each time point (TCID_50_ mL^−1^); *C*_*0*_ is the initial concentration of viable IAV PR8 measured at day 0 (TCID_50_ mL^−1^); *β*_*0*_ is the intercept; *k*_*infectivity*_ is the first-order decay rate constant for viable IAV PR8 (day^−1^); *t* is the incubation time (day).

The decay rate constant of IAV PR8 RNA (*k*_*RNA*_)was estimated by fitting a first-order decay model to the corresponding viral RNA data following equation (2):

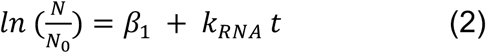

where *N* is the average concentration of IAV PR8 RNA measured at each time point (gene copies mL^−1^); *N*_*0*_ is the initial concentration of IAV PR8 RNA measured at day 0 (gene copies mL^−1^); *β*_*1*_ is the intercept; *k*_*RNA*_ is the first-order decay rate constant for IAV PR8 RNA (day^−1^); *t* is the incubation time (day).

*k*_*infectivity*_ and *k*_*RNA*_, as well as their standard error, were determined using the function “lm” in R (version 4.2.2) and Rstudio (Version 2022.12.0+353), and the goodness of fit of the first-order decay models was assessed through examination of the coefficient of determination (R^2^).

#### Pasteurization experiment

Pasteurization process is designed to inactivate the pathogens in food products and one of the most common methods used for milk pasteurization is heating the milk to 63 °C for at least 30 minutes.^18^ In this study, the raw milk spiked with IAV PR8 was heat in a water bath at 63 °C for 30 minutes to mimic the milk pasteurization process. Three independent tubes were used as experimental triplicates. 60 µL of sample was collected before and after heating and proceeded to RNA extraction immediately. The degradation of IAV PR8 RNA during the pasteurization process was determined by comparing the IAV PR8 RNA concentration before and after treatment.

#### TCID_50_ for virus titration

TCID_50_ assay quantifies the viable virus by determining the highest dilution of the sample that can cause 50% infection in cell culture.

#### Nucleic acid extraction

Extraction was performed using a ZymoBIOMICS DNA/RNA miniprep kit (#R2002, Zymo Research). Sixty microliters of milk sample were collected from the 3 replicates at each timepoint and combined with 750 μL of DNA/RNA shield (#R1100-250, Zymo Research). Samples were lysed through agitation on a Vortex Genie 2 at maximum speed for 15 minutes (#SI-0236, Scientific Industries). Negative extraction controls were included consisting of 750 μL of DNA/RNA shield and 60 μL of ultrapure water and extraction of 60 μL of raw milk which did not have spiked influenza. Positive controls were included by extracting 10 µL of IAV PR8 stock. Total nucleic acid purification was then performed and samples were eluted into 100 μL of DNAse/RNAse free water. Nucleic acid quality and concentration were assessed through spectrophotometry (#ND-1000, ThermoScientific).

#### Viral quantification

Viral RNA quantification was performed using singleplex reverse transcription digital droplet polymerase chain reaction (RT-ddPCR) using an AutoDG Automated Droplet Generator (Bio-Rad, Hercules, CA), Bio-Rad C1000 Touch Thermal (Bio-Rad) thermocycler, and a QX200 Droplet Reader (Bio-Rad). An influenza A primer/probe set targeting the M1 gene was used, consisting of CAA GAC CAA TCY TGT CAC CTC TGA C for forward primer, GCA TTY TGG ACA AAV CGT CTA CG for reverse primer, and 5’-/FAM/TGC AGT CCT /ZEN/ CGC TCA CTG GGC ACG/3IABkFQ/-3 for the probe. The probe used a FAM, 6-fluorescein amidite as the fluorescent molecule with an internal ZEN quencher (proprietary from IDT) and a 3’ Iowa Black FQ quencher.^30^ Neat cultured Influenza A PR8 was used as a positive control to validate assay performance, and no template controls were included in all runs. All extracts were diluted 100x before quantification to avoid detector saturation. Each of the triplicate extractions were run in individual wells.Thresholds on the FAM channel were set at 2,700 relative fluorescence units (RFU). Though no plates failed QA/QC, in the case of a plate failing QA/QC (i.e., positive controls and negative controls not positive and negative, respectively), we would have reported the reason for the failure and noted the re-run of any samples. Thermocycling conditions were as follows: reverse transcription at 50°C for 60 minutes, enzyme activation at 95°C for 5 minutes, 40 cycles of denaturation at 95°C for 30 seconds and annealing and extension at 61°C for 30 seconds, enzyme deactivation at 98°C for 10 minutes then an indefinite hold at 4°C.

## Results and discussion

### Persistence of influenza A viral infectivity in raw milk

The persistence of influenza viruses in raw milk was evaluated by monitoring the infectivity of IAV PR8 incubated with raw cow milk, collected from a dairy farm, at 4°C for up to 5 days. As shown in **Figure 1**, IAV PR8 decayed to below 5.0 TCID_50_ mL^−1^ after 5 days in raw milk at 4°C, with an initial concentration of approximately 4.98 × 10^5^ TCID_50_ mL^−1^. The decay rate constant *k*_*infectivity*_ was estimated by fitting a first-order decay model and was −2.05 ± 0.45 day^−1^ (estimated value ± standard error, p<0.05, R^2^=0.865). This value is greater compared to those found for influenza A virus decay in other aqueous matrices; a systematic review of enveloped virus decay in water found mean influenza virus decay to be −0.3 day^−1^ across all tested conditions (n= 560).^31^

**Figure 1.**
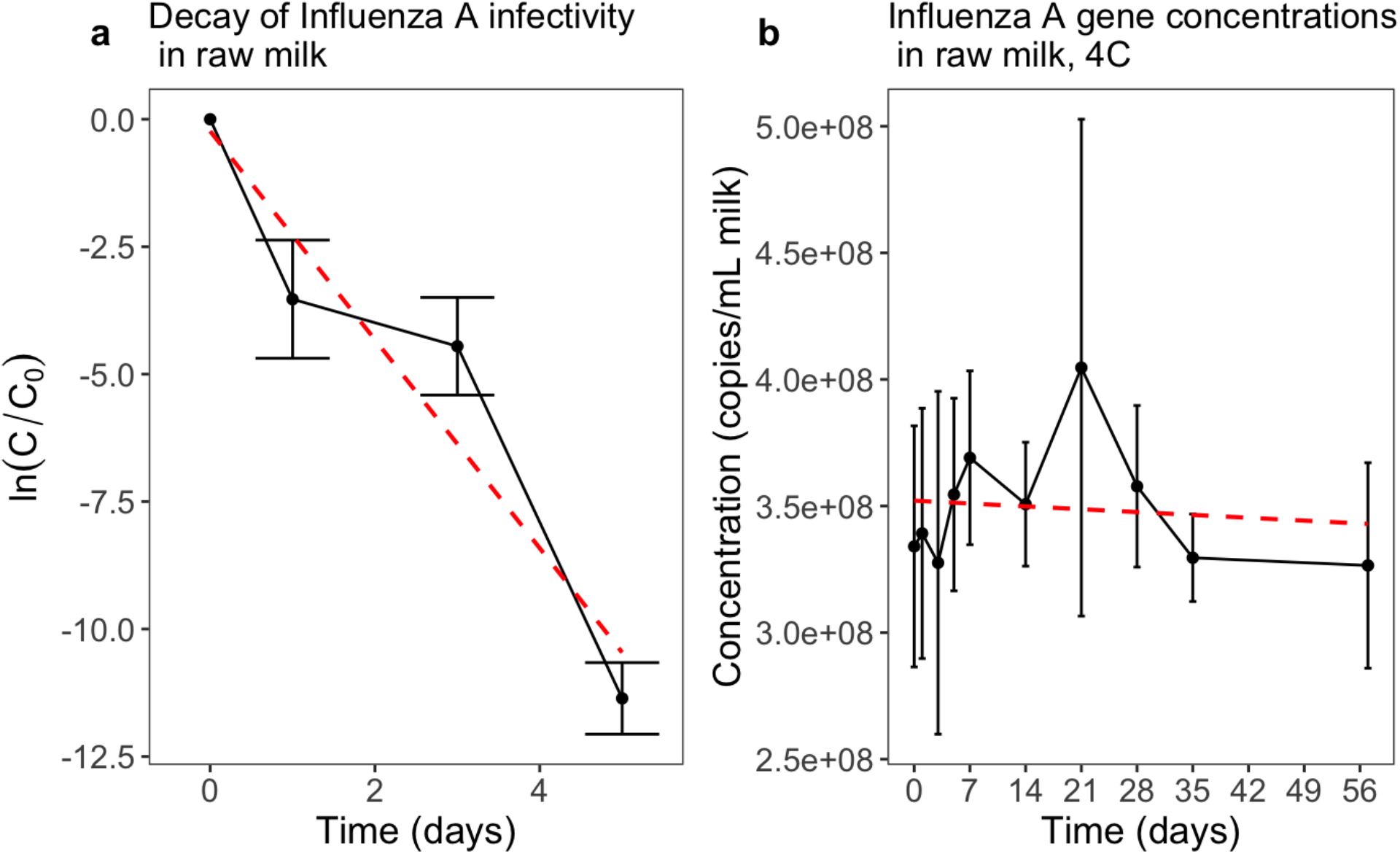
Decay of infectivity of IAV PR8 as measured by the natural logarithm of the fraction of viable IAV PR8 compared to initial time point assessed by TCID_50_ in raw milk at 4°C (1a), and decay of IAV PR8 genome copies in raw milk at 4°C (1b). The red dashed lines represent linear regression used to calculate decay rate constants. Error bars represent standard deviation based on 3 experimental replicates.

The persistence of influenza viruses in raw milk causes concern, as the consumption of raw milk remains high in the United States due to cultural factors and several popular misconceptions.^22,32^ Some of these misconceptions include beliefs that raw milk could cure lactose intolerance or asthma, enhance the immune system, and have greater nutritional value compared to pasteurized milk.^33–36^ Hundreds of outbreaks and deaths have occurred because of the pathogens contained in raw milk.^23^ Previously, only bacteria were reported in raw milk-associated disease outbreaks including campylobacteriosis, salmonellosis, tuberculosis, and *Escherichia coli* 0157:H7 infection.^21,37–39^ The results of this study show influenza A virus may remain infectious in raw milk for a prolonged period of time under refrigerated conditions, representing an potential exposure route.

Highly pathogenic avian influenza H5N1 clade 2.3.4.4b (HPAI) has been a cause for concern worldwide due to its newly observed ability to infect dairy cattle herds throughout the United States, with 243 herds in 14 states affected as of October 6, 2024.^15,40–42^ HPAI H5N1 also has a propensity to replicate in the mammary gland of dairy cattle, due to the high expression of HPAI H5N1 receptors, ultimately resulting in the shedding of HPAI H5N1 in the milk.^17^ Mammal-to-mammal transmission has been observed in the form of cats ingesting raw milk on dairy farms, and subsequently being infected with HPAI, further supporting that HPAI H5N1 can be shed into milk.^43^ Le Sage et al. reported that influenza H5N1 and H1N1 remained infectious in unpasteurized milk on stainless steel and rubber surfaces for up to 5 hours.^20^ The data presented here significantly contributes to existing knowledge by demonstrating that influenza A can remain infectious in raw milk for up to 5 days at 4°C. This shows that there could be significant risk of inter-species transmission of the virus due to environmental spillage of milk. The recommended shelf life of raw milk under refrigeration is 7-10 days.^44^ Based on our results, it took 2.3 days to achieve 99% reduction in infectivity (T99) of IAV PR8 in raw cow milk (Figure S1).

One limitation of this study is that we measured the decay of exogenous human IAV spiked into raw cow milk. The decay of endogenous HPAI H5N1 in the milk from HPAI H5N1–infected dairy cattle could potentially be affected by presence of antibodies in milk. It is known that the influenza-specific antibodies in human breast milk can provide protective immunological properties for the infants and play an essential role in increasing resistance of infants to influenza virus infection.^45^ However, little is known about the impact of antibodies present in milk on the persistence of influenza viruses shed in milk, which call for further research.

### Persistence of influenza A viral RNA in raw milk

IAV PR8 nucleic acids remained detectable and stable for 57 days in raw milk at 4°C (Figure 1). The calculated decay rate constant *k*_*RNA*_ is −0.00049 (p=0.709). This value is not significantly different from 0. Further, we took samples from time point 0 and pasteurized them as described above. The resulting pasteurized milk samples were extracted and IAV RNA quantified using the same protocol as the raw milk. The resulting ddPCR concentrations showed a significant reduction from an average of 2.9 * 10^8^ copies/mL in the raw milk samples to 9.1 * 10^7^ copies/mL in the pasteurized milk samples, a decrease of 70% (Fig 2, p<0.01, Welch’s t-test).

**Figure 2.**
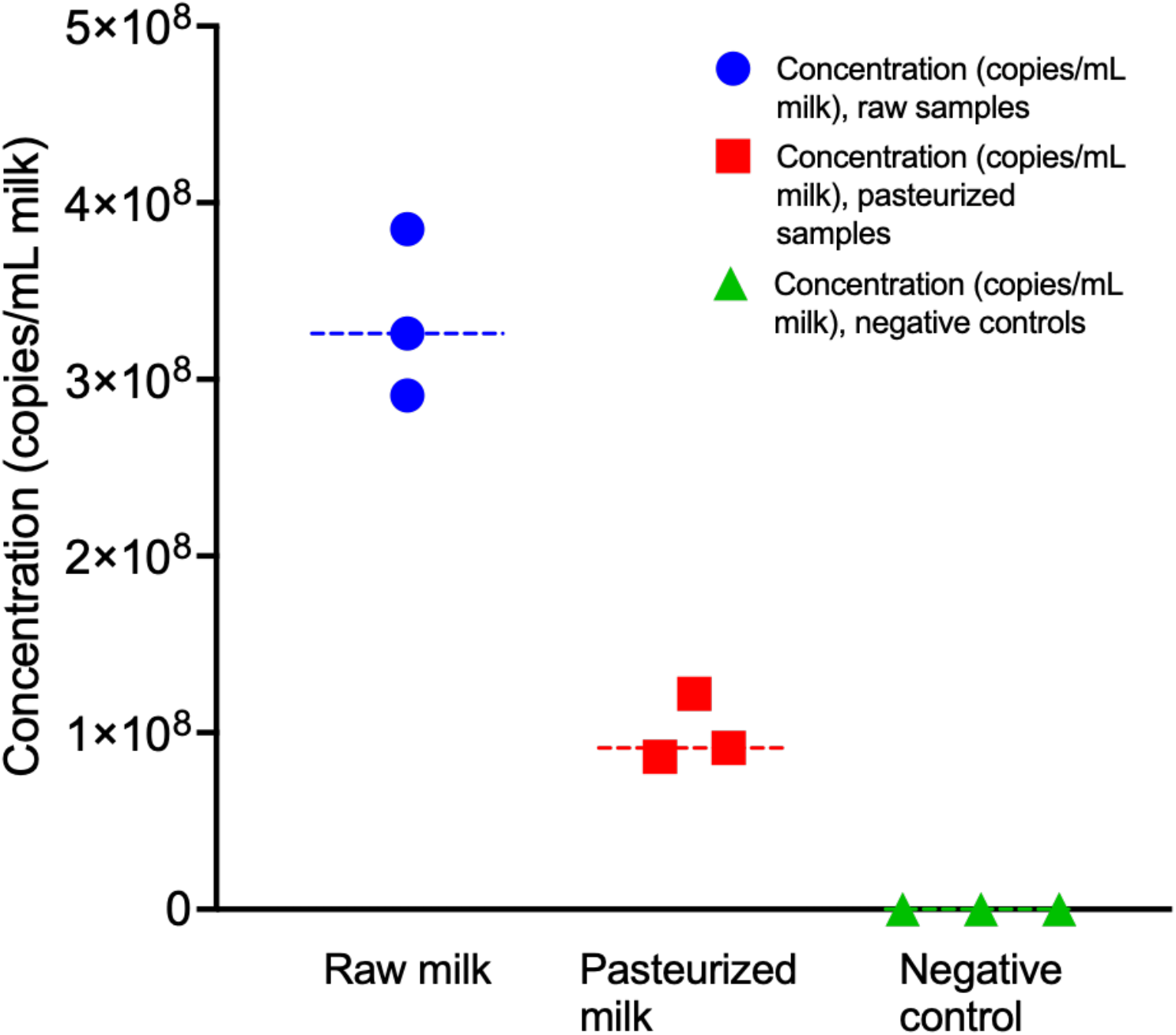
Measured concentrations (in gene copies per ml of milk) of influenza in raw milk samples (blue), pasteurized samples (red), and negative no template controls (green). The horizontal dashed lines represent the mean of the data points. All negative controls were found to be negative.

HPAI has been extensively detected in wastewater streams across the United States, and the source of these detections was determined to most likely have occurred due to industrial discharge from local milk processing facilities.^27,46^ However, inputs from residential or commercial discarded milk are also possible sources.^46^ It is estimated that 17 kg of milk and dairy products are discarded per capita per year by consumers, translating to 15,500 tons per day.^47^ Given our findings of the stability of influenza A RNA in milk, residential or commercial milk discarded through drains could represent a source of H5N1 RNA in wastewater. This high level of stability potentially indicates that environmental spillage of milk could also lead to detections within the environment.

Our study has significant implications for both environmental and food safety. The extremely prolonged persistence of IAV PR8 RNA in raw milk, compared to that of virus infectivity, may complicate efforts to assess the safety of raw milk and other dairy products. This is because the detection of viral genomic materials, a detection method that is rapid and relatively easy to implement, may not necessarily indicate the presence of viable viruses. Pasteurization has been shown to successfully inactivate influenza viruses, including HPAI, in milk.^24^ However, HPAI has been consistently detected through molecular biology methods in pasteurized milk from affected cattle herds,^48^ which could be explained by the high resistance of IAV RNA to pasteurization reported herein. Moreover, source tracking might be essential when interpreting wastewater surveillance data for influenza viruses; industrial discharges may be a source of H5N1, but so could household or commercial milk discarding, and also potentially inputs from other sources including infected humans or birds.^27^

Overall, our study demonstrates that influenza viruses remain infectious in raw milk for up to 5 days, which could pose a significant human health risk, and concentrations measured through molecular biology methods remain constant for at least 57 days. The results presented here will help both public health officials understand the risks of raw milk consumption, and provide context for environmental scientists and engineers working in the environmental detection of influenza. It is important to note that this study used H1N1 influenza A subtype because H5N1 is a select agent and working with it in a laboratory requires extensive safety protocols and certifications. Results of experiments using other subtypes could vary. However, a systematic review of the literature on influenza A decay in water suggested that decay of most subtypes was similar in that matrix.^31^

## Data Availability

All data produced in the present work are contained in the manuscript

## Notes

### Competing Interest Statement

The authors have declared no competing interest.

### Funding Statement

This study was supported by Woods Institute for the Environment at Stanford University and by the Sergey Brin Family Foundation.

