## Supporting Information for "Infectivity and persistence of influenza viruses in raw milk"

### Methods

#### Details of virus propagation.

MDCK cells were maintained in Dulbecco's modified Eagle's medium (DMEM) supplemented with 10% fetal bovine serum (FBS), 1% Penicillin-Streptomycin (P/S), and 2 mM L-glutamine and incubated at 37℃/5% $CO_2$ until confluence. The confluent MDCK cells were rinsed twice with Phosphate-buffered saline (PBS) buffer (Fisher Scientific, MT21040CV, pre-sterilized) and once with cDMEM/0.2%BSA before inoculation with IAV PR8, since the FBS inhibits viral entry and must be removed for efficient infection. cDMEM/0.2%BSA is the DMEM medium supplemented with 1% P/S, 2 mM L-glutamine, 25 mM HEPES buffer, and 0.2% bovine serum albumin (BSA). IAV PR8 stock was diluted with influenza virus growth medium, which was cDMEM/0.2%BSA supplemented with 2 µg $mL^{-1}$ L-1-Tosylamide-2-phenylethyl chloromethyl ketone (TPCK)-treated trypsin, and inoculated into the confluent MDCK cells at a multiplicity of infection (MOI) of 0.1. Infected cells were incubated at 37℃/5% $CO_2$ until significant cytopathic effect (CPE) was observed, which usually took 2-3 days. Virus suspension was collected and proceeded to further purification and concentration.

#### Details of TCID50

Briefly, the collected samples were serially diluted 10-fold with influenza virus growth medium and inoculated into confluent MDCK cells, which were maintained in 96-well plates and rinsed twice with cDMEM/0.2%BSA before inoculation. The inoculum was removed after 3 hours incubation to ensure virus binding and replaced with fresh influenza virus growth medium. Five replicate wells were used for each dilution. The 96-well plates were continually incubated at 37℃/5% $CO_2$ for 3 days, when significant cytopathic effect (CPE) was observed. After the supernatant was removed, the cells were stained with crystal violet in 70% ethanol for 10 minutes, which also inactivated the viruses. All the wells that were transparent or had large clearings in the cell lawn were counted as positive and the concentration of viable IAV PR8 was calculated as $TCID_{50}$ $mL^{-1}$ following the Spearman–Kärber method. The lower detection limit (LoD) was defined as the concentration where only one out of five wells was scored as positive at the lowest dilution ($10^{-1}$), corresponding to 5.0 $TCID_{50}$ $mL^{-1}$ in this study. Five wells inoculated with 10-fold diluted raw milk were used as negative control to rule out contamination from the milk. The Penicillin-Streptomycin in the influenza virus growth medium would prevent the growth of bacteria in raw milk that were added onto the cells.

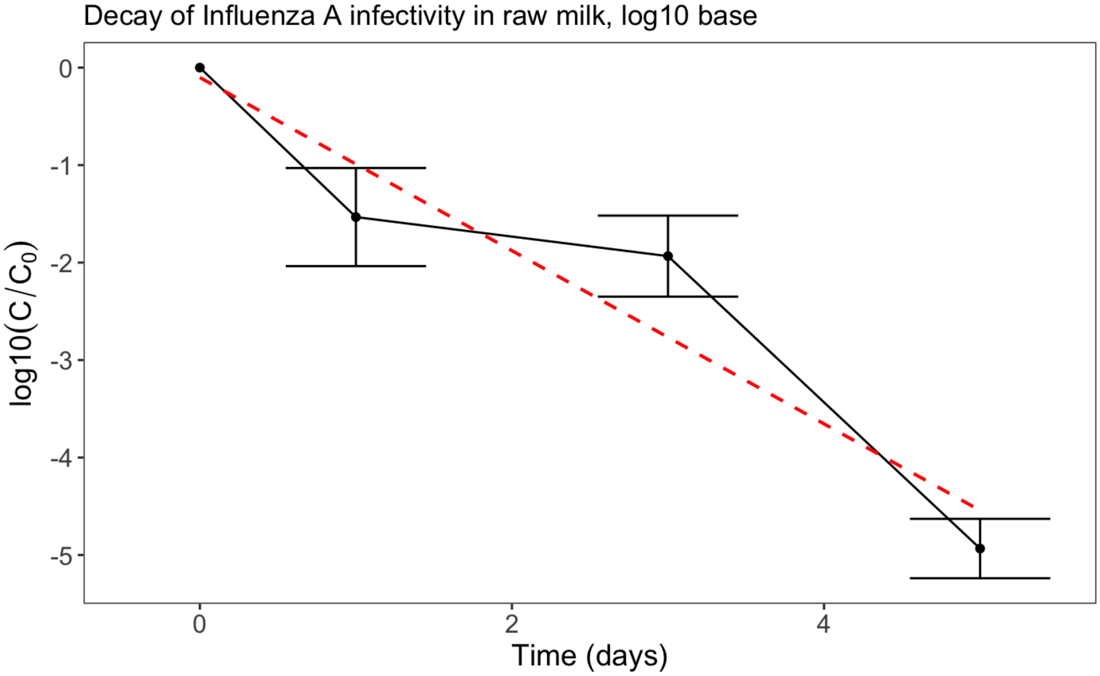


Figure S1: Decay of infectivity of IAV PR8 as measured by the logarithm-base 10 of the fraction of viable IAV PR8 compared to initial time point assessed by $TCID_{50}$ in raw milk at 4°C. The red line represents a linear regression. The error bars represent standard deviations.
